# Perception of yips among professional Japanese golfers: perspectives and challenges

**DOI:** 10.1101/2021.02.18.21252043

**Authors:** Gajanan S. Revankar, Yuta Kajiyama, Yasufumi Gon, Issei Ogasawara, Noriaki Hattori, Tomohito Nakano, Sadahito Kawamura, Ken Nakata, Hideki Mochizuki

**Author notes:** **Correspondence**: Hideki Mochizuki, Department of Neurology, Graduate School of Medicine, Osaka University, 2-2, Yamadaoka, Suita, Osaka, Japan – 5650871, Ken Nakata, Department of Health and Sport Sciences, Graduate School of Medicine, Osaka University, 2-2, Yamadaoka, Suita, Osaka, Japan – 5650871. **Financial disclosure related to research**: None.

## Abstract

**Background:** Yips in golf is a complex spectrum of psychological anxiety and movement disorder that affects competitive sporting performance. Existing literature is limited to several western studies and the manifestations of this problem in Japanese golfers is currently unknown.

**Objectives:** To quantify self-reported perception and manifestation of yips among Japanese golfers from the professional golfers’ association (PGA).

**Methods:** We analyzed 1271 (of 1356) elite golfers in a cross-sectional manner. Golfers were sensitized beforehand about yips by a movement-disorder specialist. Based on a positive history for yips, participants were categorized into yips and non-yips groups. Survey questionnaire focused on demographic information, golfing habits, anxiety and musculoskeletal problems, performance deficits during golfing, changes in training and their outcomes. Statistical procedures included multiple logistic regression and network analysis to assess factors associated with yips.

**Results:** 35% (N=450) of the respondents had experienced symptoms of yips in their career, their odds increasing proportionally to their golfing experience. Severity of musculoskeletal symptoms were higher in those with yips. Regardless, about 57% of all yips-golfers attributed their symptoms to psychological causes. Putting, approach and teeing shots, in that order, were highly susceptible to movement problems. Network analysis highlighted characteristic movement patterns i.e. slowing, forceful or freezing of movement for putting, approach and teeing respectively. Golfers’ self-administered strategies to relieve yips symptoms were generally inconsequential, though improvements were seen only for approach-yips.

**Conclusion:** Our findings align firmly with prior studies on yips. Though aware of the problem, most Japanese golfers were untouched by yips. Those that were affected, perceived yips to be a psychological issue despite substantial evidence pointing to a movement-disorder. While self-administered interventions for symptom relief in such golfers is satisfactory at best, it may be imperative to sensitize golfers from a movement-disorder standpoint for early identification and management of the problem.

## Introduction

Yips in professional golfers presents as sudden, acute loss of motor skill resulting in loss of performance during high-pressure sporting environments. Often, this is attributed to situational stress or competition anxiety though substantial evidence with respect to muscle contractions and movement kinematics suggest it to be a task-specific movement disorder^1–3^. Not limited only to golf, the yips has been known to affect other sports as well, some of which include cricket, baseball, archery and snooker^4^.

In yips, the anxiety component generally triggers as ‘choking’ wherein athletes fail to execute an action due to their perception of attentional resources as being insufficient^5^. Fundamentally, this relates to how sports anxiety is defined i.e. an unpleasant state of stress resulting in under-performance^6^. As a result, yips is considered to be a neuropsychological phenomenon due to its congruity with high-pressure situations and its near-complete absence of symptoms in day-to-day activities. However, this argument more or less overlaps with how task-specific dystonias are defined as well. Task-specific dystonia are involuntary excessive muscle contractions in repeatedly learnt skilled activity occurring specifically during its performance^7,8^. While the pathophysiological aspects of task-specific dystonia are still under exploration, lately, the yips phenomenon has been used synonymously under the umbrella term of focal dystonias^9^.

Considering the strong psychiatric comorbidities associated with task-specific dystonias^10^, this duality in yips may be a lot harder to distinguish without causal evidence. Prior efforts have provided substantial anecdotal evidence, predictors, mechanisms and theories but a clear etiology is still elusive^4^. Several well-designed experimental work and case-reports have addressed crucial psychogenic components or movement-disorder related aspects of yips^1,3,11,12^. On the other hand, studies have also employed questionnaires or semi-structured interviews to evaluate the causative symptoms of yips^13–15^. Notably, Smith et. al. performed a focused investigation among yips-only golfers to subjectively evaluate their ‘perception’ regarding yips^16^. The rationale behind this exercise was to observe the influence of self-perception on their symptoms, their golfing habits and their compliance towards getting professional support (psychological, neurological or orthopedic) with respect to the performance deficit. This study, and several others focusing on western populations^12,15,17^, provided a promising approach to triage yips symptoms via participant derived responses to facilitate management of such golfers.

Equipped with the above information, we analyzed a large-scale cross-sectional survey among golfers in Japan, the groundwork formulated previously on highly skilled professional and amateur golfers^18^, to ascertain their demographic characteristics, golfing habits, their experience with yips and the kinematic issues associated with it. Additionally, we investigated what strategies golfers implemented to relieve their symptoms of yips and the outcomes of such measures. Our aim of this study was to present a systematic clinico-demographic description of yips in Japanese professional golfers, literature of which is currently sparse. Furthermore, beyond the qualitative analysis, we also exploited network methodologies to analyze the survey dataset since network structures provide an efficient modality to examine interactions between large number of variables^19,20^. Prior studies have highlighted the potential advantages of incorporating logistic regression through network analysis in neuropsychiatric disease datasets to demonstrate the relationships between different symptoms^21–23^.

## Methods

### Design and population

We surveyed participants from the Professional Golfers’ Association (PGA), Japan, focusing specifically on elite golfers with a professional teaching license. The study was performed between 2016 and 2018 wherein golfing information was collected from respondents aged 18 and above. Participants were recruited via in-person seminar sessions and training workshops that were held in PGA. The paper-based survey questionnaires were handed out to a total of 1356 participants. Prior to its distribution, a movement-disorder specialist sensitized the golfers regarding the features, characteristics and the spectrum of yips. The Osaka University institutional review board (IRB) for clinical research approved this study. Informed consent was obtained from all the golfers who participated in the survey.

### Survey measures and outcomes

Demographic and golfing details in the questionnaire included age, gender, years of golfing experience, handedness, practice hours per month, number of golfing rounds per year and number of private practice rounds. Questions related to musculoskeletal problems were provided with multiple responses with a severity scale that encompassed movement problems during competitions (low), during practice and competitive golf (moderate), and those that affected activities of daily living (severe). Information regarding presence of anxiety or nervousness in public was requested to evaluate trait-based characteristics. Based on a single question that asked whether the golfers suffered from yips or not, additional questions followed. Such of those who responded positively for presence of yips, details regarding their speculated cause of yips (a movement disorder, or a psychological disorder or something else), their problem type related to club use (wood, iron, wedge or putter) and problems in shot type (tee, fairway, driver, rough, bunker, approach and putt) was obtained. To define the movement problems of the golfers, we documented them under a pre-defined list of five most common types, since feature definitions from free-response questions have been shown to be extremely diverse in yips golfers^16^.

Accordingly, these included forceful shots, sluggish/slowing of shots, tremors, jerks and freezing. Lastly, participants were requested to provide details of their strategies to relieve the symptoms of yips that listed changes in golfing techniques (e.g. grip changes, use of gloves, changing length/size of the clubs, handedness, etc. i.e. free-responses), increasing or decreasing the frequency of training (practice hours), and the outcome of these changes (improved, worsened or no change). Based on this formulation, we reported the differences between normal and yips golfers and an exploratory analysis of the influence of such variables among yips only group.

### Statistical analysis

Descriptive statistics were reported as mean and standard deviations (SD) for relevant demographic variables. Missing values for demographic data were imputed using nearest neighbor method (knnimpute function on Matlab). Missing values counts were, for age N=1, for experience N=2, for practice hours N=30, and for rounds/year N=38. To tests the differences between the normal and yips group, non-parametric Mann-Whitney test was performed with statistical significance set to p < 0.05. In order to ascertain which of the demographic factors were associated with yips, we performed a multivariate logistic regression analysis specifying a binomial distribution representing the 2 groups (with and without yips). The proportional odds ratio and confidence intervals for the above variables were reported for p < 0.05 (Wald’s statistic). To evaluate anxiety and severity of musculoskeletal problems between the groups, a Chi-square test with alpha=0.05 was performed along with graphical representation of frequency estimates.

A subgroup analysis was performed with respect to yips only group. Frequency estimates were calculated for club-type, shot-type, movement disorder-type and training-type categories. Considering the large number of variables, we utilized network analysis to gather a holistic picture of yips golfers’ characteristics. In brief, a network comprises of several ‘nodes’ of interest and the link between these nodes are defined by its ‘edges’^24^. With respect to our dataset, each dependent variable served as a ‘node’, whereas ‘edges’ represented the relationship between the variables after conditioning with respect to other variables. Considering the categorical binary nature of our data, we applied a recently developed Ising model of logistic regression to study the network structure. Ising models have shown to be a promising approach to study large psychological datasets that are predominantly populated with ‘yes’/’no’ type data^25^. Network analysis was performed on shot-type, movement disorder-type and training-type using IsingFit function on JASP statistical software. Club-type estimates were excluded due to high collinearity with shot-type data. Edge parameters rules included: non-zero regression coefficients between nodes (‘AND’ rule) and gamma hyperparameter value at 0.25. To ascertain the accuracy of edge-weights^26^, we bootstrapped the edges of the estimated network and their centrality on 1000 random networks. Between-node edge weight confidence intervals which did not overlap with other non-zero nodes were considered significant. Nodes in the network were positioned via the Fruchterman-Reingold algorithm wherein the structure was defined based on the nodal connection strength^27^. When reporting the effects between different outcomes, network structures were constrained for clear visualization and comparability. Subsequent network properties were then analyzed with respect to degree centrality, betweenness and expected influence of nodal properties^28^. Degree centrality defined the importance of the variable in the network, and was illustrated by the number of connections of that node to all other nodes in the graph^29^. Betweenness centrality provided a quantification of the node serving as a bridge between two other connected nodes along its shortest path. Finally, in a network with positive and negative edges, the expected influence metric factored in the negative associations among the nodes for interpretations on the variable importance^28^.

All preprocessing of data was performed on Matlab v2018b (MathWorks, USA). Statistical analysis and graphing were done on JASP software (v0.14) that uses relevant R packages (qgraph) for network analysis. For statistical tests, p-values lower than 0.05 were considered significant. Data analyzed in this manuscript will be made available from the corresponding author upon reasonable request.

## Results

Demographics characteristics of the respondents are described in Table-1. We obtained 1271(out of 1356) participant responses from the questionnaire achieving a 94% response rate for the survey.

**Table-1:**
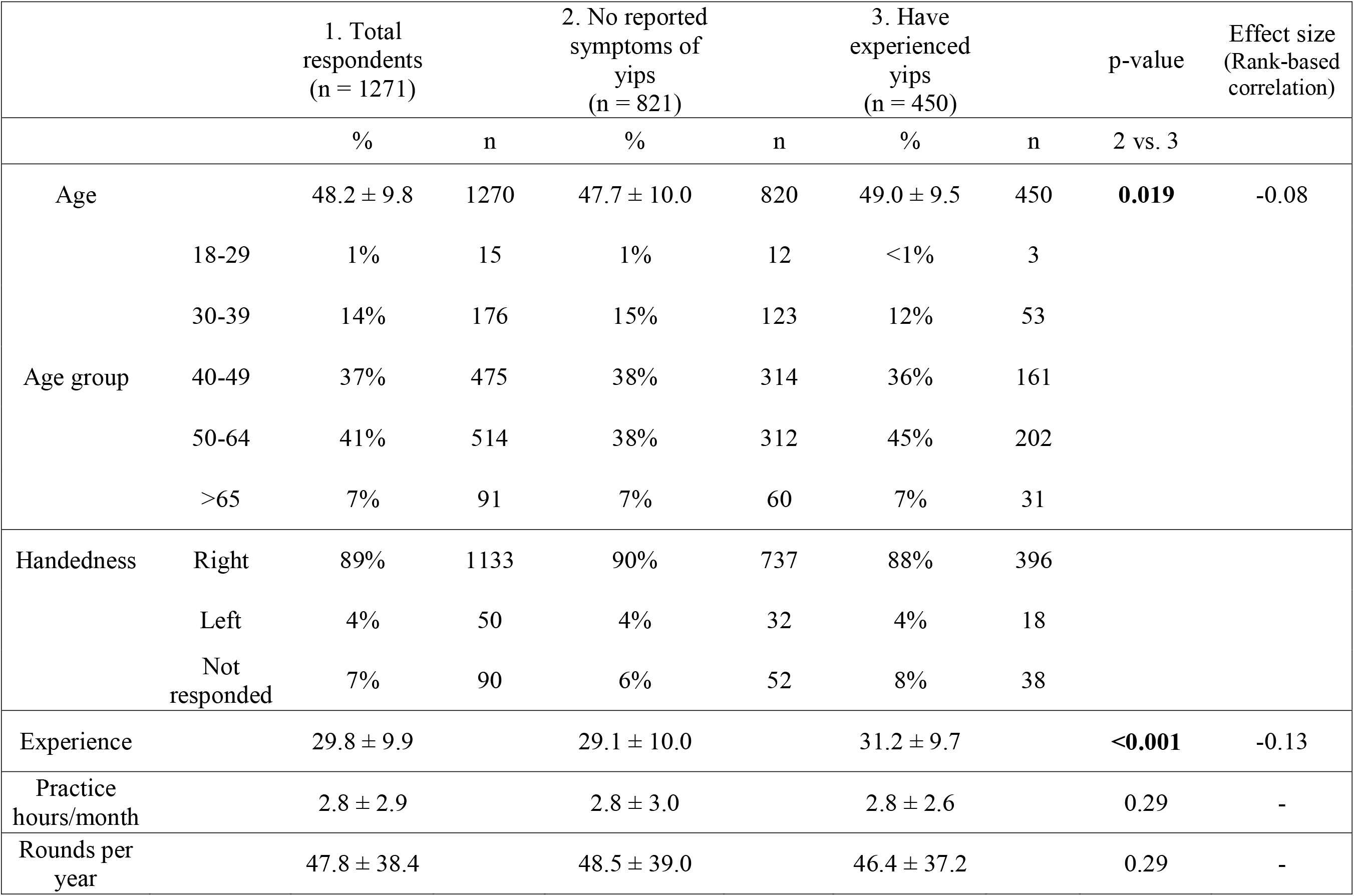
Demographic characteristics of professional golfers. Reported p-values are derived from Mann-Whitney non-parametric test.

About 35% of the respondents professed to have had yips during their career (N=450). A majority of such yips golfers (approx. 45%) were in the 50 to 64 year age category. Age and experience among the participant groups, though statistically significant, were roughly similar (effect sizes approx. 0.1, rank-based correlation).

Multivariate logistic regression analysis of the binomial variable ‘Group’, given the reference level of all other factors, showed golfing experience to be a significant predictor in the occurrence of yips (OR, CI = 1.043, 1.02-1.07, p < 0.001), Figure-1.

**Figure-1:**
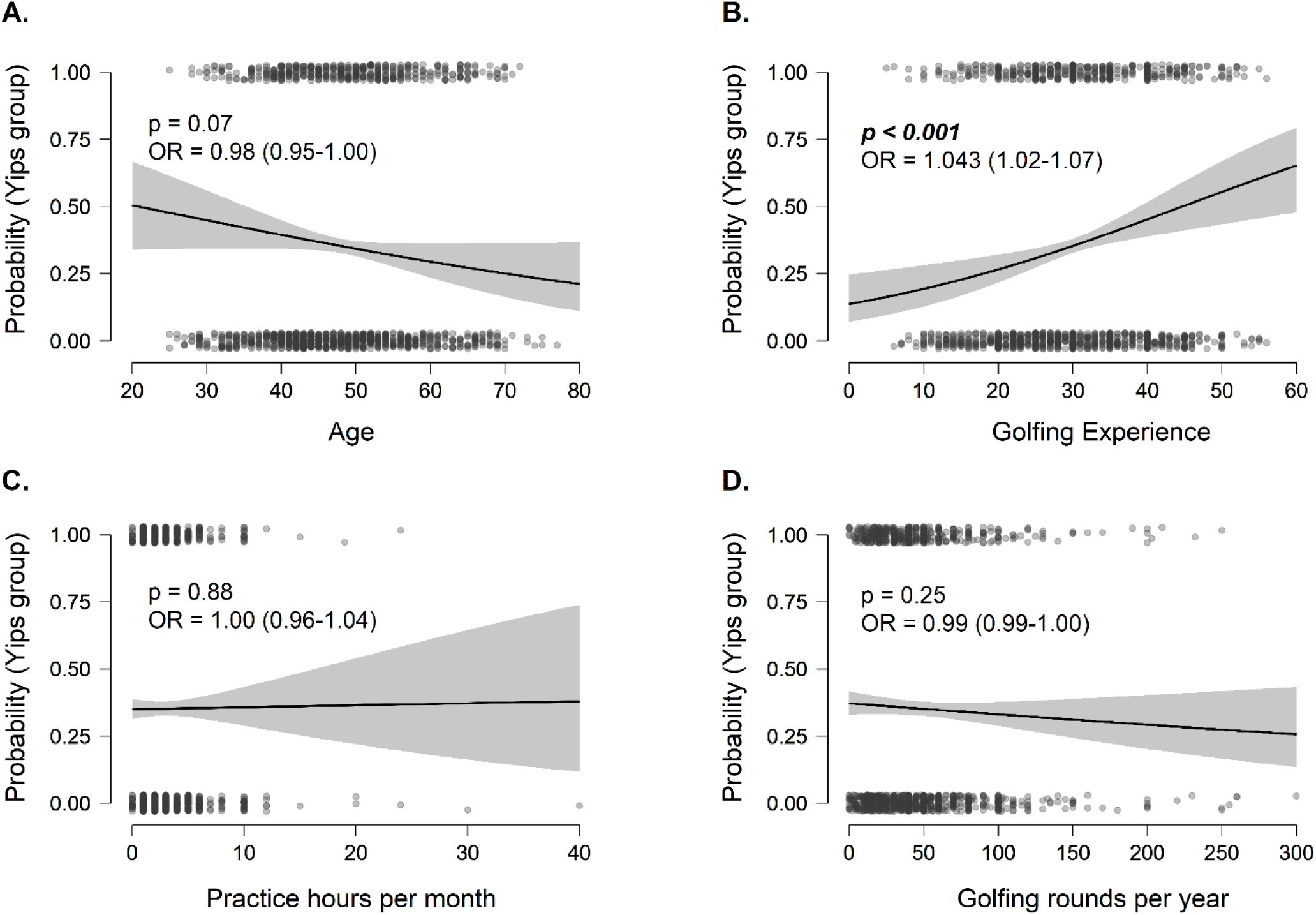
Multiple logistic regression of demographic predictors for yips. Conditional estimates plot for A. Age, B. Golfing experience, C. Practice hours/month and D. Rounds/year. OR = Odds ratio with confidence intervals for odds ratio. Yips groups coded as 1. Golfers with yips: N=450, Golfers without yips: N=821.

More golfers with yips answered that their personality was of nervous disposition (proportional test, chi-square test (χ^2^) = 20.25, p<0.001). The responses for the possibility of at least a single musculoskeletal injury were similar between the groups (χ^2^ = 2.46, p=0.117). However, yips golfers had a higher proportion of severe musculoskeletal problems (χ^2^ = 7.60, p=0.006). When asked what the golfers felt their yips was due to, nearly 57% of yips golfers attributed the performance deficits to psychological causes. Only a meagre 5% felt yips was due to a movement-disorder, Figure-2.

**Figure-2:**
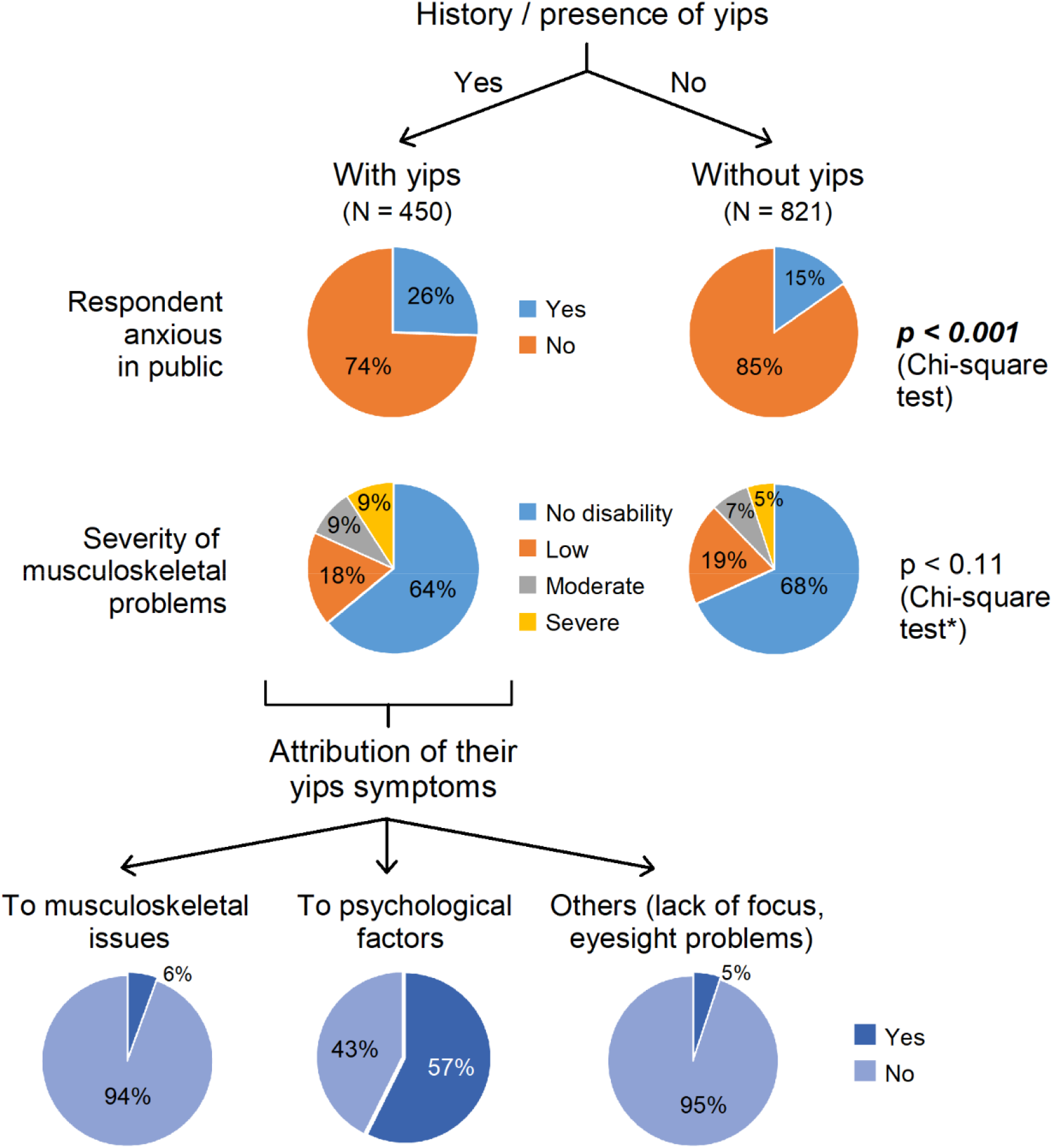
Response frequencies of golfers’ perception of yips. Perception and attribution of symptoms among respondents. *Chi-square test represents the grading of at least one musculoskeletal problem (grades = none, low, moderate, severe) among the golfers. Significant differences were observed between normal and yips golfers having severe musculoskeletal problems (χ2 = 7.60, p=0.006).

For subgroup analysis among yips-only respondents (N=450), 29 golfers were excluded for one or more of the following reasons: (i) stopped playing golf altogether, (ii) did not specify any current symptomatology due to yips, or (iii) erroneous entries or unanswered questions; bringing the total number of yips-only respondents to N = 421. The problems with their use of club, the type of shots, the underlying movement deficit associated with it and any accompanying interventions for relief of symptoms are summarized in Figure-3.

**Figure-3:**
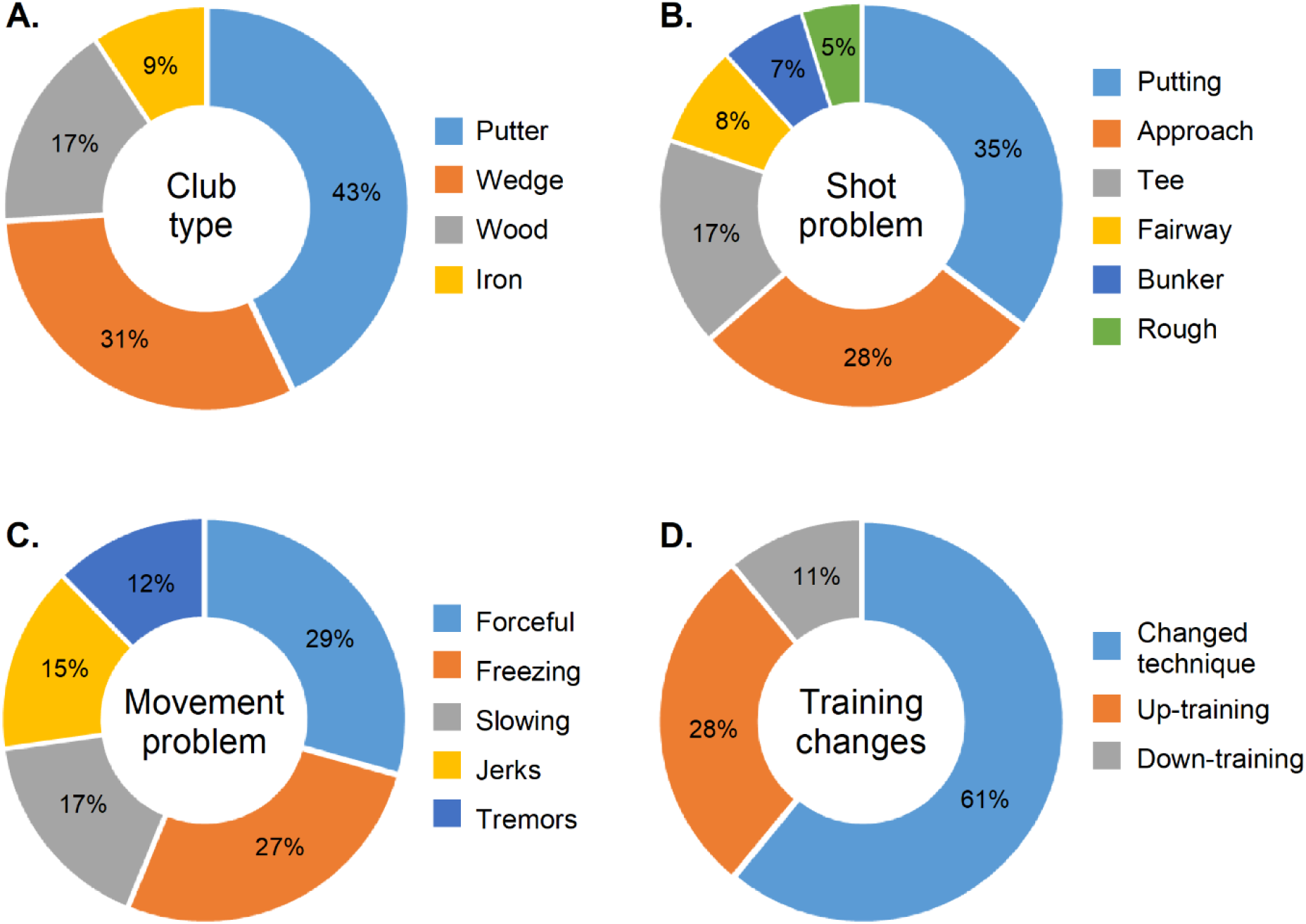
Frequency estimates of problem types and interventions in yips only golfers (N=421)

The above variables were then grouped for network analysis to study their relationships. For shot-type, we focused on putting, approach and tee shots since they comprised the dominant types of shot problems (approx. 80%). Subsequent network estimation revealed the following key features (Figure-4): (i) a strong positive regression between putting and slowing of shots; between approach and forceful shots; and between tee shots and freezing of movement, (ii) a negative relationship between changing technique following yips and increasing the frequency of training, (iii) high degree centrality and influence of putting shots suggesting linkage between several other problematic movement-types, and (iv) high degree centrality and influence for changing technique and up-training compared to down-training for most golfers.

**Figure-4:**
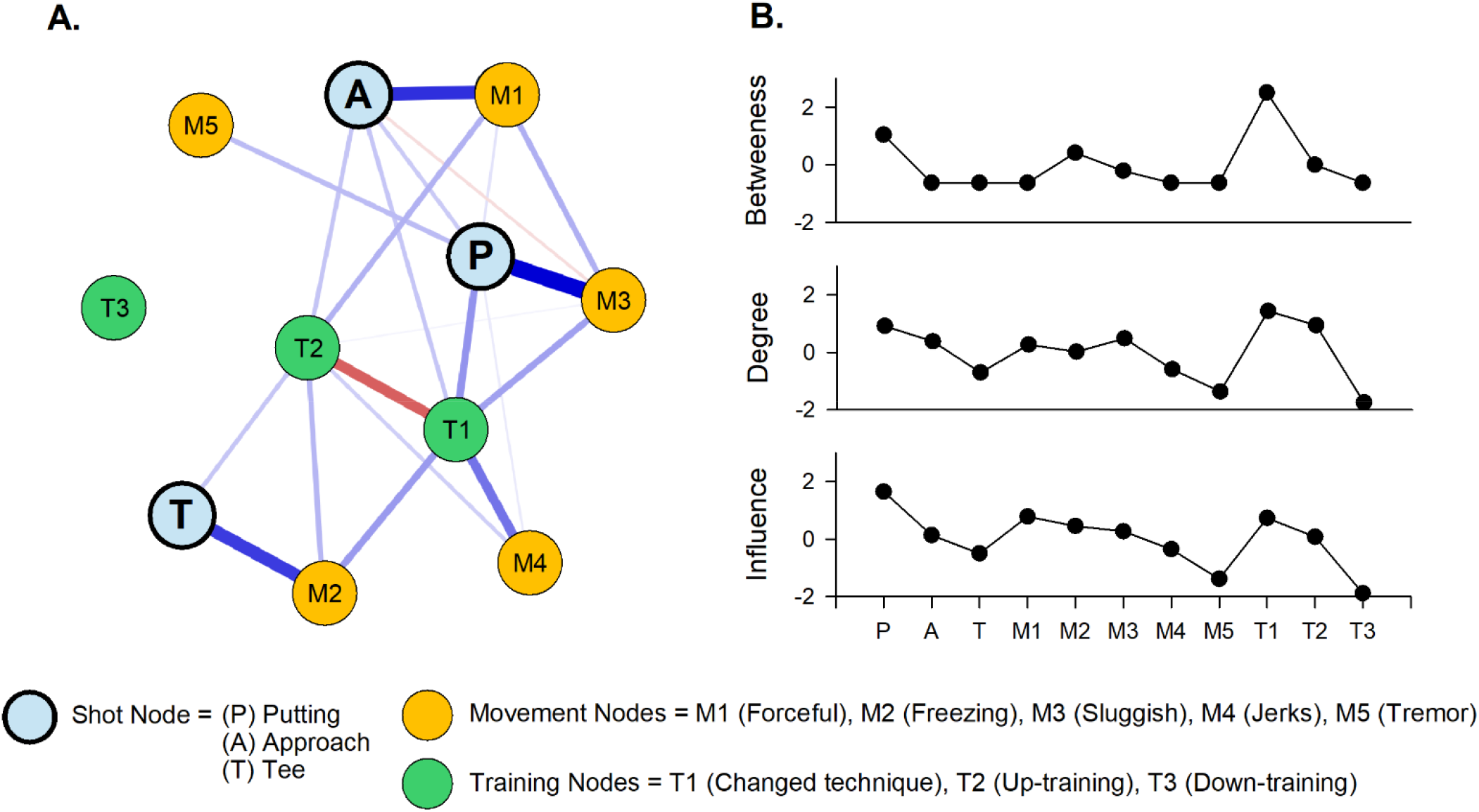
Network and centrality plot of shot-type, movement problems and interventions to relieve yips symptoms. A. Network structure for three variables (nodes) = shot-type, movement-type and training type. Network edges were based on an Ising model (hyperparameter = 0.25) wherein blue edges represent positive relationship between the variables and red edges represent negative relationships. The strength of the connection between the nodes was proportional to the thickness of the edges represented between them. Only non-zero edges are shown, with sparsity = 0.56. The distance between the nodes was arbitrary, specified by a value given by the Fruchterman-Reingold algorithm (here = 0.7). The edge-weight stability demonstrating the robustness of the network is summarized in supplementary data. As seen from the network, putting(P), approach(A) and tee shots(T) nodes had strong edge connection to specific movement deficits (slowing, forceful and freezing of swing, respectively) in the network. B. Centrality plot with respect to the network structure was explained via betweenness, degree and the explained influence of the variables (nodes) in the network. The X and Y axes specify the nodes and the indices respectively. Putting shots along with their predominant training patterns i.e. changing technique and increasing the frequency of training had strong degree centrality with significant influence on yips-golfers’ characteristics within the network.

We then evaluated golfers’ strategies to counter the yips symptoms. Of the 421 respondents, only 253 answered if their techniques relieved (N=102), worsened (N=11) or showed no change (N=140) in their symptoms. For network analysis, we excluded golfers whose symptoms worsened due to very low sample sizes. Nevertheless, network structures were extremely sparse, Figure-5 (Sparsity index=0.83).

**Figure-5:**
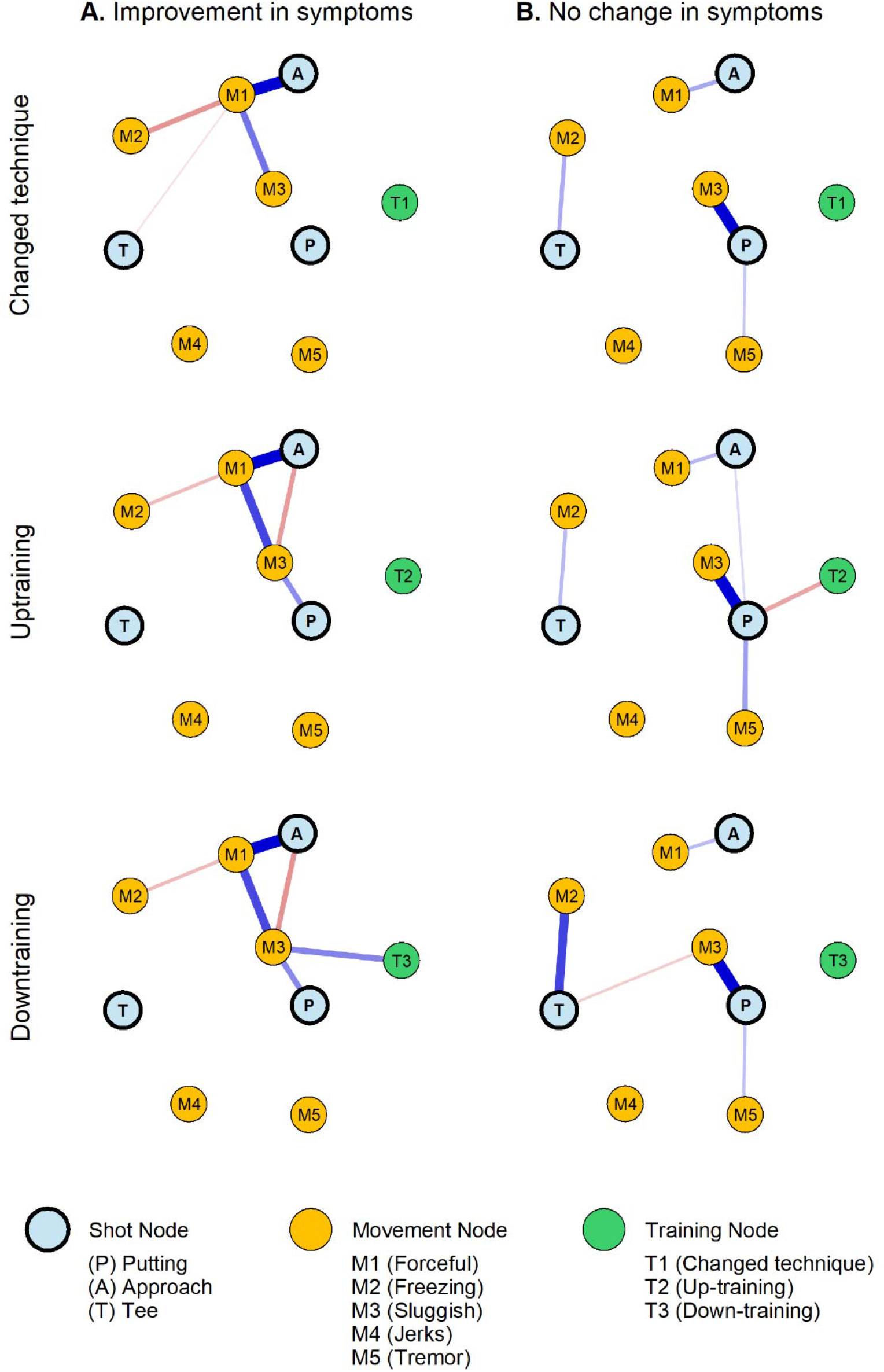
Network plots of outcome of interventions to relieve yips symptoms. In comparison to Figure-4, the network plots were specified for each strategy that the golfers responded as: A. An improvement in their symptoms or B. No change in their symptoms. Due to the low sample sizes, the network construction was different from Figure-4 in that (i) Golfers with worsening of symptoms were omitted, (ii) non-zero regression coefficients were specified with ‘OR’ rule, (iii) hyperparameter γ=0, (iv) Fruchterman-Reingold distance=1, and (v) network sparsity = 0.83. Network representation were constrained to a fixed ‘X’ and ‘Y’ axis coordinate system for visual comparison purposes. The resulting network showed that approach shots were more pliable for changes in self-administered training adjustments. On the other hand, golfers who reportedly did not show any change in symptoms were mostly with putting type of yips.

For each strategy type, golfers’ perception of improvement was seen only for approach shots. Those who reported no change in symptoms showed weaker connections between shot and the movement types, with putting shots being impervious to any improvements.

## Discussion

With a sizable response rate, our survey report provides a comprehensive overview of the perception of yips among Japanese golfers and quantifies the burden associated with it. In golfers who had the yips, we observed that (i) long golfing experience plays a crucial role in precipitating the symptoms of yips, (ii) kinematically, putting, approach and tee shots are frequently affected, in that order, each accompanied by a characteristic movement deficit, and (iii) whereas approach yips seems receptive to improvements via adjustments in their techniques or training patterns, the same does not hold true for putting or teeing yips in a majority of such golfers. Our findings also hint that despite objectively reporting problems in the musculoskeletal system, a substantial number of yips-golfers perceive yips to be a psychological issue. This perception begets self-help strategies that appear futile in the management of yips without professional help.

Our cohort conformed several well-established findings observed in yips. Notably, the high proportion of yips golfers among older adults (50 to 60 age group), a mean prevalence of yips that hovers around 30% (among Western populace), the probability of yips occurrence with long golfing exposure and the high predisposition to anxiety in such golfers, are in line with previously documented studies^4,13,17^.

Additionally, within our group, we observed that nearly 57% of yips-golfers attributed their symptoms to be psychological. We speculate that this perception among golfers may play a key role in creating a degree of anxiety that may impinge upon their gameplay. Models that explain sports related anxiety conceptualize that the cognitive self-evaluation and stress response if left unchecked, result in increased muscle tension, loss of focus and attention along with a range of other physiological behavioral changes^6^. This means that up to a certain point depending on the individual’s own threshold of sense of anxiety, the performance-anxiety loop can either streamline their quality of shots or can potentially debilitate the task^30^. Anxiety tests should therefore serve as a baseline in any yips assessment with every golfer requiring the necessary appraisal to improve their performance. On the other end of the yips spectrum are task-dependent dystonias identified as involuntary excessive muscle contractions in repeatedly learnt skilled activity^8,12^. Despite the overarching presence of anxiety in our survey, we observed that yips golfers had significantly higher musculoskeletal symptoms that affected not only their golfing performance, but also activities outside of their competitive environments.

While traditionally, this does not fit into the definition of task-specific dystonia, one possible explanation for these findings are that these could be the same muscles that are required for executing day-to-day tasks. Given that symptoms of task-specific dystonia precipitate when there is continuous, intensive over-use of specific musculature for long periods of time^31^, these golfers are likely to be susceptible to movement disorders with or without performance anxiety^7^.

Professional golfers often spend considerable practice time in perfecting the putting stroke^32^. Not surprisingly, we found that putting was the most affected stroke in yips-golfers, a finding that has been definitively reported in other studies as well^1,2,17^. The high degree centrality, noted via network analysis, showed that putting was accompanied by a variety of movement problems that resulted in abnormal control of swing i.e. forceful or slowed motion of the swing, or those suggestive of abnormal co-contractions i.e. tremors, jerks or freezing. Of the above, slowing of movement during putting was the most frequent. Approach and tee shots were characterized by forceful-strikes and freezing respectively. In light of the low degree and betweenness centrality for approach and tee shots, the above-mentioned movement problems for these types may be deemed specific.

The inclusion of training type within this network revealed certain important findings. First, both changing technique as well as up-training (increasing the frequency) had high degree centrality (positive edges to all movement-types) suggesting that golfers frequently tried one of these interventions. Second, the strong negative relationship between the two implied that these strategies were mutually exclusive. Those who up-trained avoided changing their technique, a strategy seen frequently for approach or teeing yips. However, changing technique outweighed other modalities and considering its relationship with diverse muscular symptoms for putter’s yips, our findings reinforces the magnitude of performance deficits that golfers face in ameliorating putting symptoms.

Down-training showed no definitive relationship with other variables. Down-training, along with retraining, have shown favorable outcomes with respect to dystonic symptomatology^8,33–36^. Statistically, our findings may be a direct effect of low sample sizes since the network construction in Ising models are critically dependent on it^25^. Still, it is vital to recognize why only about 10% of our respondents lowered their training habits. With high attribution rates (∼57%) of yips to psychological causes, we speculate that this may simply be due to a (mis)conceptualization of the movement problem. Regardless, those who ascribe yips to a ‘choking’ phenomenon or perceive their crisis due to lack of practice are both at risk of overtraining and contributing to the abnormal plasticity^37^. It would therefore be necessary to sensitize the golfers about the benefits of rest and immobilization in rehabilitative retuning of the symptoms.

With the above self-administered strategies in place, network structures revealed that irrespective of the type of intervention, golfers’ perception of improvement was higher for approach shots than with putting or tee shots. Approach shots, in general, constitute the broadest category in golf in terms of distance from the putt^38^ and without causal evidence it may be difficult to conclude why such adaptability was seen among our golfers. On the other hand, golfers with putter’s yips formed the biggest group who showed no changes in their symptoms. We reason this could be a consequence of the diverse movement problems reported for putting shots. However, we wish to provide a conservative viewpoint for these outcomes since the networks were extremely sparse (along with bootstrapped confidence-interval overlaps), lowering the interpretations of the network structure.

### Limitations

It is important to consider that only a few of the respondents (N=6) from the survey had received a formal clinical diagnosis of task-specific dystonia. While the term ‘yips’ is relatively common among golfing circles, the knowledge of yips rested on the participant via seminars offered by the movement-disorder specialists. We were therefore careful in interpreting our findings without excessive speculation, since most respondents were not formally/physically diagnosed by a movement-disorder specialist. In the current scenario, with a plethora of information available over the internet, it is likely some respondents may have overestimated their problem. Contrarily, some of the symptoms may have been underreported due to a level of stigmata attached to the problem among athletes. Despite these subjective intricacies, we reckon that the golfers were well-informed about yips and therefore their responses were reasonable. In a functional movement disorder such as yips, we believe our methodology of assessment was in-line with prior studies with the sampling variations to be fair. Furthermore, the survey detailed several movement-related problems in yips inadvertently creating a bias wherein anxiety issues may not have been appropriately assessed. Addressing these aspects is likely to improve the quality of assessment in the future thereby allowing longitudinal comparisons as well as opening up to encompass athletes from other sports as well.

## Conclusions

Yips in golf has a stigma attached to it is that is often problematic^16^. Whether the cause is anxiety or dystonia, the outcome is unitary – a distinct, appreciable loss in performance wherein mild cases may not be troublesome but as severity increases may essentially disrupt athletes’ careers. Fortunately, a majority of professional Japanese golfers are unaffected by the yips. While our report highlighted the characteristics, features and similarities with several other prior studies on yips, there were some observations among Japanese golfers that does necessitate deliberations. Despite the musculoskeletal and/or kinematic issues, a majority of yips-golfers speculate their condition to be psychological. With no apparent benefits owing to training changes, it may be crucial to target golfers early in their career with an assertive informational outreach regarding the movement-disorder aspect of yips. For health-care providers, it will be imperative to address these challenges to influence golfers’ compliance towards accessing timely remedies for their symptoms.

## Supporting information

STROBE_Reporting guidelines

## Data Availability

Data that support the findings of this study will be made available from the corresponding author upon reasonable request.

## Funding

This work was supported by the Sports Research Innovation Project (SRIP) grant, sponsored by the Japan Sports Agency.

## Contributions

G.S.R., Y.G., Y.K., N.H., I.O., S.K., K.N. and H.M. conceptualized and designed the study. N.H., Y.G., Y.K. and T.N. disseminated the questionnaire and collected data. G.S.R. and Y.K. analyzed the data and performed the statistical analysis. G.S.R wrote the manuscript. All authors reviewed the manuscript, suggested corrections and approved its final version.

## Notes

**Conflict of interests**: None.

### Competing Interest Statement

The authors have declared no competing interest.

### Author Declarations

The Osaka University institutional review board (IRB) for clinical research approved this study.

