## Supplementary data for "Perception of yips among professional Japanese golfers: perspectives and challenges"

**Supplementary Figure-1: Edge weight stability**

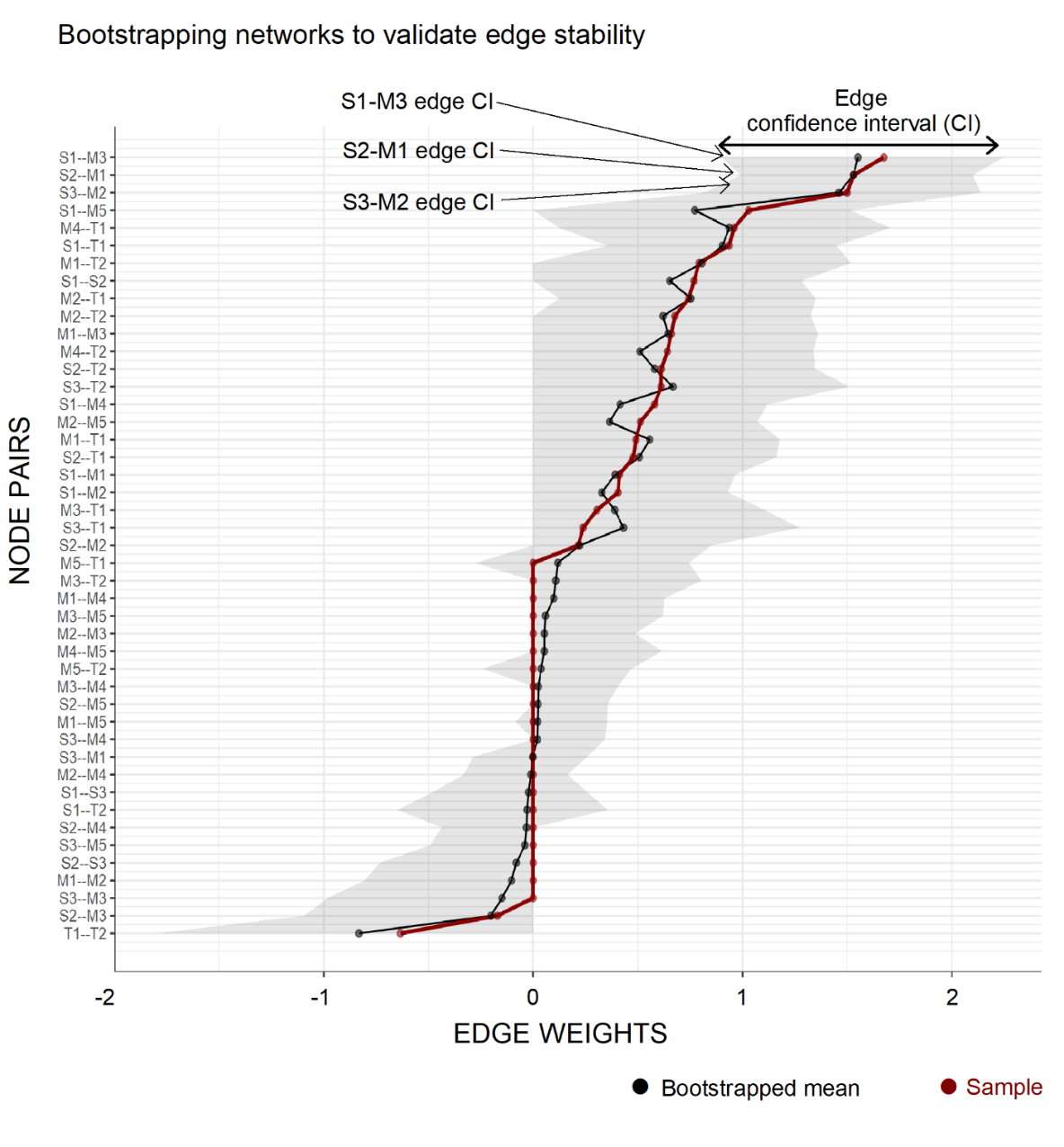

*Supplementary Figure-1 caption: Graph shows between-nodes edge weight confidence interval (CI) for sample and bootstrapped means. Red scatter-line plots represent original edge weight of the network. Black scatter-line plot denotes bootstrapped edge weights of random networks. The random networks were created by bootstrapping 1000 times. Edge CI which did not overlap with other nodes of interest were considered significant. Notably, edge CI for putting (S1), approach (S2) and tee shots (S3) pairs with their corresponding musculoskeletal problems (M3, M1 and M2 respectively) were significantly different with other shot-movement problem pairs.*
